# Aberrant Cortical-Subcortical-Cerebellar Connectivity in Resting-State fMRI as an Imaging Marker of Schizophrenia and Psychosis: A Systematic Review of Data-Driven Whole-Brain Functional Connectivity Analyses

**DOI:** 10.1101/2025.06.19.25329865

**Authors:** Kyle M. Jensen, Tricia Z. King, Pablo Andrés-Camazón, Vince D. Calhoun, Armin Iraji

## Abstract

Schizophrenia is extremely heterogenous, and the underlying brain mechanisms are not fully understood. Many attempts have been made to substantiate and delineate the relationship between schizophrenia and the brain through unbiased exploratory investigations of resting-state functional magnetic resonance imaging (rs-fMRI). The results of numerous data-driven rs-fMRI studies have converged in support of the disconnection hypothesis framework, reporting aberrant connectivity in cortical-subcortical-cerebellar circuitry. However, this model is vague and underspecified, encompassing a vast array of findings across studies. It is necessary to further refine this model to identify consistent patterns and establish stable imaging markers of schizophrenia and psychosis. The organizational structure of the NeuroMark atlas is especially well-equipped for describing functional units derived through independent component analysis (ICA) and uniting findings across studies utilizing data-driven whole-brain functional connectivity (FC) to characterize schizophrenia and psychosis. Towards this goal, a systematic literature review was conducted on primary empirical articles published in English in peer-reviewed journals between January 2019 - February 2025 which utilized cortical-subcortical-cerebellar terminology to describe schizophrenia-control comparisons of whole-brain FC in human rs-fMRI. The electronic databases utilized included Google scholar, PubMed, and APA PsycInfo, and search terms included (“schizophrenia” OR “psychosis”) AND “resting-state fMRI” AND (“cortical-subcortical-cerebellar” OR “cerebello-thalamo-cortical”). 10 studies were identified and NeuroMark nomenclature was utilized to describe findings within a common reference space. The most consistent patterns included cerebellar-thalamic hypoconnectivity, cerebellar-cortical (sensorimotor & insular-temporal) hyperconnectivity, subcortical (basal ganglia & thalamic) – cortical (sensorimotor, temporoparietal, insular-temporal, occipitotemporal, & occipital) hyperconnectivity, and cortical-cortical (insular-temporal & occipitotemporal) hypoconnectivity. Patterns implicating prefrontal cortex are largely inconsistent across studies and may not be effective targets for establishing stable imaging markers based on static FC in rs-fMRI. Instead, adapting new analytical strategies, or focusing on nodes in the cerebellum, thalamus, and primary motor and sensory cortex may prove to be a more effective approach.

---

Schizophrenia is a severe psychiatric disorder and major cause of disability worldwide (Theodoridou & Rössler, 2010). While much progress has been made over the last few decades to establish the biological profile of schizophrenia and understand the underlying neural mechanisms (Dabiri et al., 2022; Meyer-Lindenberg, 2010), much work is still needed to establish stable imaging markers with clinical applications (Insel & Cuthbert, 2015; Jablensky, 2010; Morris et al., 2022). The clinical presentation of schizophrenia is highly heterogenous, with a great amount of variability across individuals (Wolfers et al., 2018), although psychosis (e.g., hallucinations, delusions, and disorganized behavior and speech) is generally considered the most characteristic feature, and is consequently the focus of much research.

## Aberrant Cortical-Subcortical-Cerebellar Connectivity

Resting state functional magnetic resonance imaging (rs-fMRI) is a neuroimaging method that has proven to be useful for identifying markers of schizophrenia as it avoids bias linked to specific tasks and is non-invasive and less intensive than modalities involving experimental tasks, making it ideal for clinical populations (Fu et al., 2024). These approaches have experienced rapid growth in recent years, reaching a record high in 2019 with more than 100 peer-reviewed articles published utilizing rs-fMRI to examine schizophrenia populations (Fu et al., 2024). However, like individual symptom profiles, the reported patterns of aberrant functional connectivity (FC) derived from rs-fMRI are also heterogenous (Jiang et al., 2013). The disconnection hypothesis of schizophrenia (Friston, 1998) has been widely applied to help interpret these findings (Friston et al., 2016), postulating that schizophrenia reflects a dysfunctional integration of neuronal activity. The specific patterns of dysconnectivity have often been characterized by Andreasen and colleagues’ (1998) theory of cognitive dysmetria which poses that symptoms of schizophrenia arise from disruptions in cortical-subcortical-cerebellar circuitry.

Although Andreasen et al. (1998) originally emphasized disruptions in circuitry between the cerebellum, thalamus, and prefrontal cortex, this framework has become somewhat of an underspecified umbrella term encompassing many different findings spanning the whole brain. For example, Matsuo et al. (2013) reports reduced activation in the inferior frontal gyrus, thalamus, and cerebellum, while Walther et al. (2017) reports hyperconnectivity between the motor cortex and thalamus, between the motor cortex and cerebellum, and between the subthalamic nucleus and anterior cingulate and dorsolateral prefrontal cortex. Yao et al. (2025) uses a slightly more specific variation of the term, “cerebello-*thalamo*-cortical”, to describe disruptions in circuitry between the cerebellum and postcentral gyrus, between the thalamus and middle temporal gyrus, and between the thalamus and middle and inferior occipital gyri. “Cerebello-*thalamo*-cortical” centers the model specifically on the thalamus and emphasizes its modulatory role (Harikumar et al., 2023; W. J. Hwang et al., 2022), however, this model still lacks specificity as the thalamus is a central hub in many brain networks (K. Hwang et al., 2017). Importantly, the findings of these three studies are largely non-overlapping, and yet they all use variations of “cortical-subcortical-cerebellar” to describe their findings. While this framework has proven to be useful in helping to overcome cortico-centric bias (Parvizi, 2009) by incorporating subcortical and cerebellar structures into pathological models of schizophrenia and psychosis, these heuristics overly generalize findings and should be supplemented with more descriptive terms, such as directionality and specific subcortical and cortical structures, if the field is to establish stable and reliable imaging markers.

## Addressing Inconsistency and Heterogeneity with NeuroMark

Perhaps due to the highly interdisciplinary nature of the field, or because it is a relatively young branch of science, a disinclination to articulate findings through clear and consistent nomenclature has been identified as a major weakness within the field of neuroscience (Uddin et al., 2019, 2023). While there are many factors potentially contributing to these inconsistencies, individual subject variability across subjects within the same study (see Figure 1 in Jensen, Turner, et al., 2024) as well as within a single subject over the course of an fMRI scan (Iraji et al., 2019) only complicates the issue further. Thus, there is great benefit in efforts towards standardization, such as those employed by the NeuroMark approach (Du et al., 2020; Iraji et al., 2023; Jensen, Turner, et al., 2024), which utilizes data-driven methods to identify functional units sensitive to subject, dataset, and study level differences and adapt them into a common reference space.

**Figure 1.**
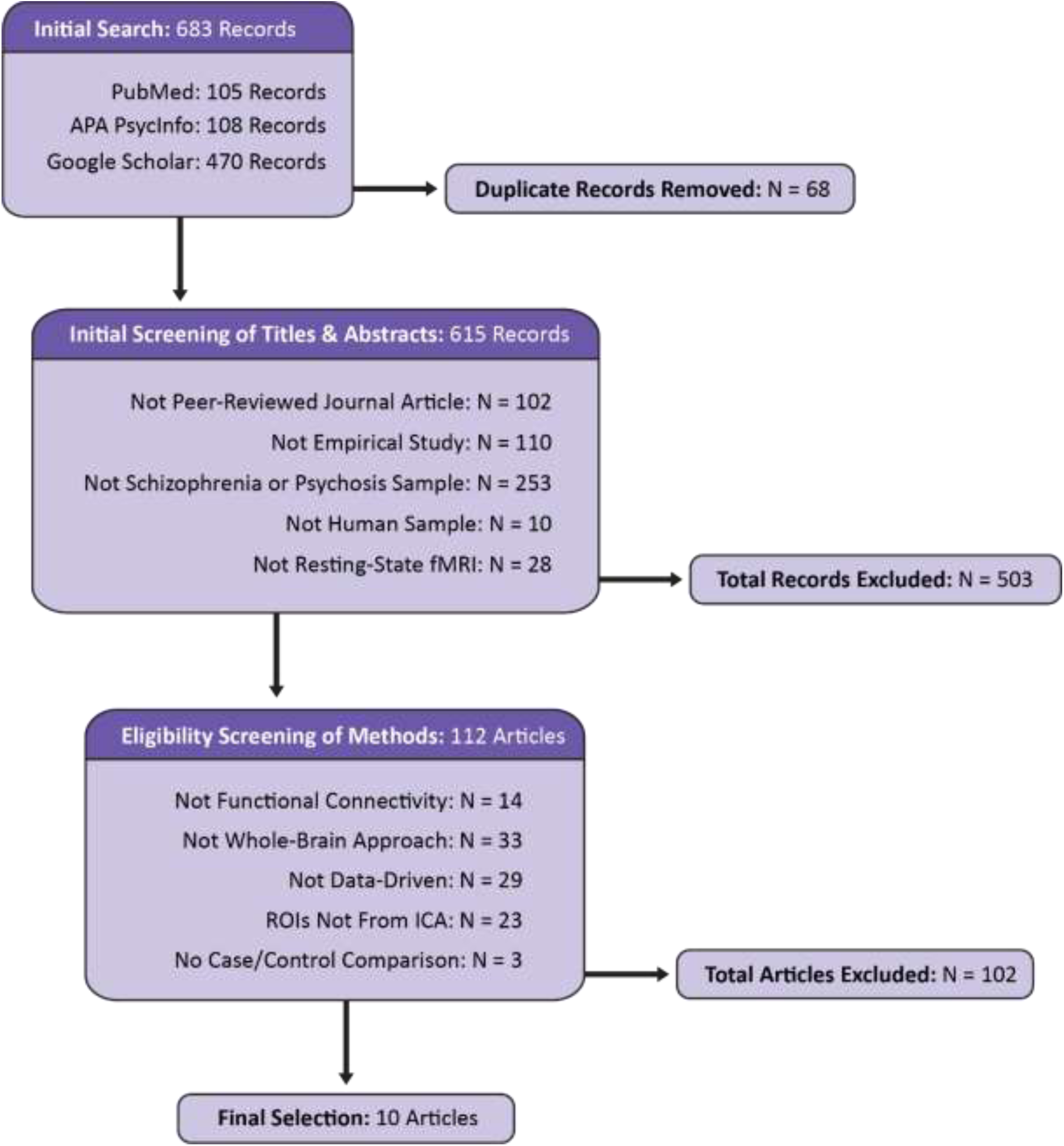
Flow diagram displaying the identification, screening, and selection of articles for the current review. Modeled after the PRISMA 2020 template (Page et al., 2021).

Whole-brain data-driven functional connectivity methods, such as those utilizing group independent component analysis (ICA; Calhoun et al., 2001) to extract intrinsic connectivity networks (ICNs), allow for unbiased exploratory approaches which yield rich and comprehensive results (Calhoun et al., 2009). However, the large amount of information produced by data-driven approaches such as these can be a double-edged sword. While they have great potential to facilitate valuable new discoveries, one of the challenges inherent with data-driven approaches lies in tasks of organizing, summarizing, and synthesizing vast amounts of information (Calhoun et al., 2021; Hutchison et al., 2013). Unfortunately, the challenge of interpreting these findings is a burden which is often placed on the reader (Allen et al., 2012). Furthermore, it can be difficult to compare findings across studies employing blind ICA due to variations in the identified ICNs (Abou-Elseoud et al., 2010; Du et al., 2020). Such inconsistencies across studies contribute to the heterogeneity currently hindering the development of a coherent biologically-informed model of schizophrenia.

Within the last five years, the NeuroMark approach has made great strides towards harmonizing results across studies by employing spatially-constrained ICA (Lin et al., 2010) to incorporate spatial priors or templates derived from large datasets (Du et al., 2020; Iraji et al., 2023). The NeuroMark 2.2 atlas has further improved the accessibility and interpretability of findings across studies by translating a previously developed data-driven template into terms familiar to the fields of cognitive and affective neuroscience (Jensen, Turner, et al., 2024). This template consists of 105 ICNs which cover the whole brain (see Table 1 and Figure 2 in Jensen, Turner, et al., 2024), incorporate information from multiple spatial scales, and have demonstrated reliability across the lifespan (Bajracharya et al., 2024).

**Figure 2.**
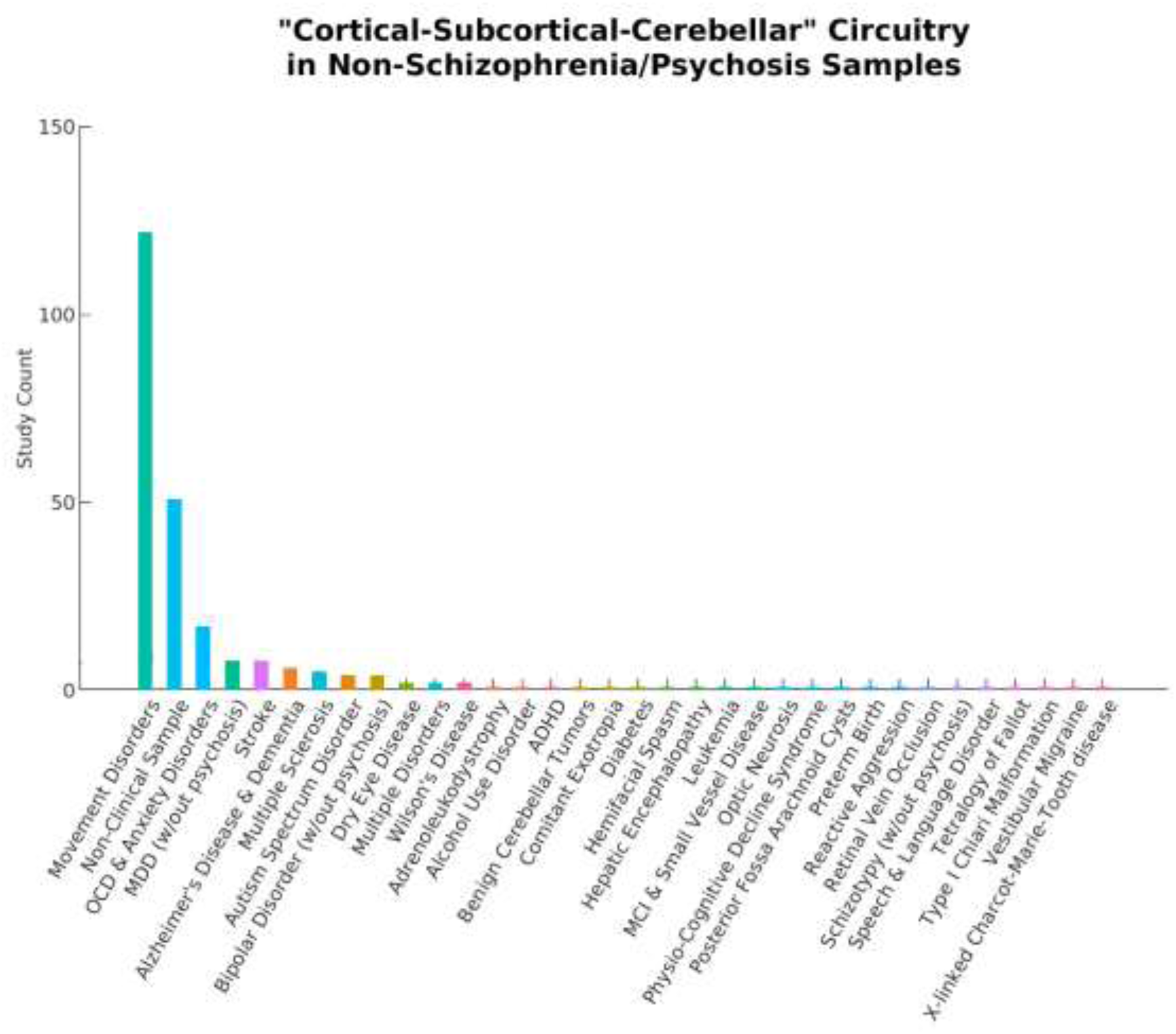
A total of 253 articles using the term “Cortical-Subcortical-Cerebellar” to describe their findings were excluded from the current literature review because they did not include schizophrenia or psychosis samples. The bar chart above displays a wide range of study samples and the number of articles for each. Movement disorders include Parkinson’s Disease, Tourette’s syndrome, tremors, dystonia, chorea, ataxia, myoclonus, epilepsy, and lateral sclerosis. OCD: Obsessive Compulsive Disorder; MDD: Major Depressive Disorder; ADHD: Attention Deficit and Hyperactivity Disorder; MCI: Mild Cognitive Impairment

**Table 1.**
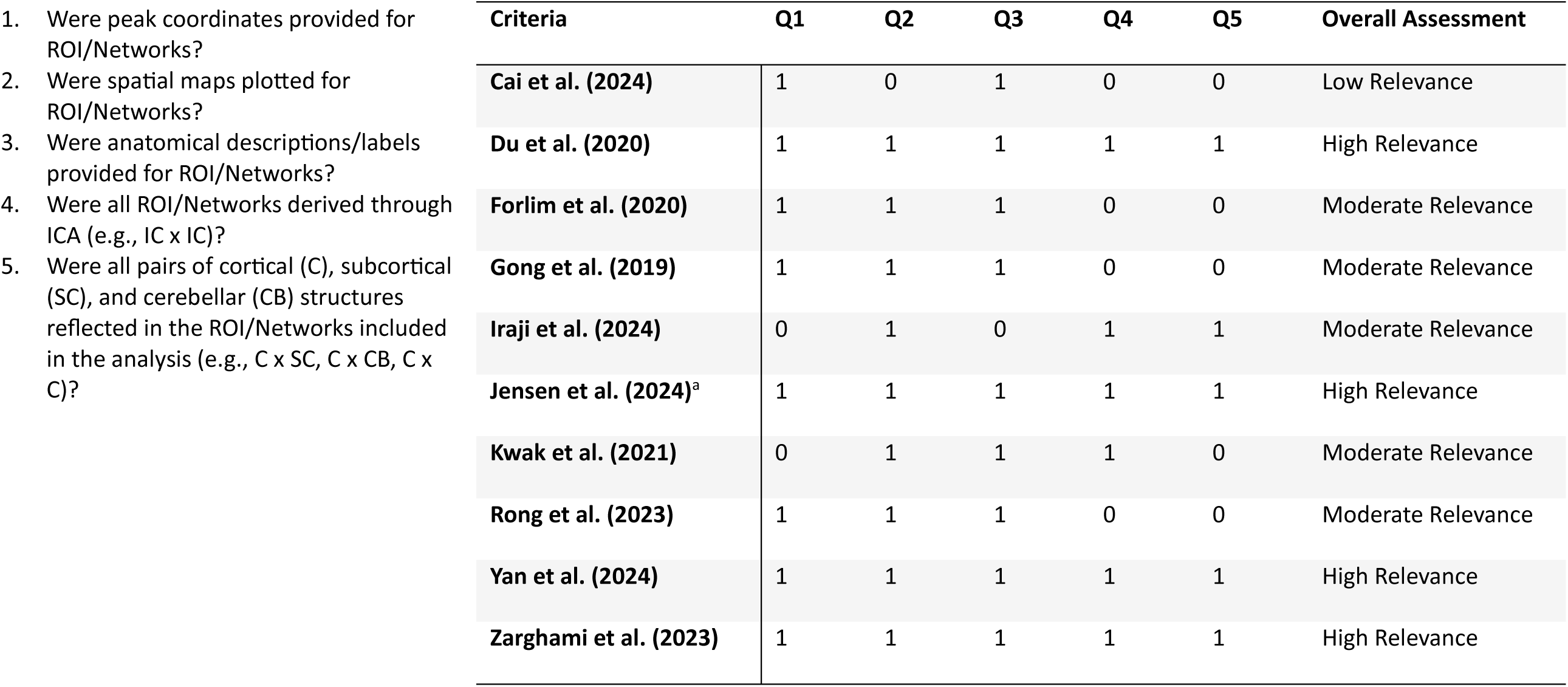
Relevance assessment. The relevance of each study included in the review was rated with the following criteria evaluating the attributes which enable study results to be accurately translated into the NeuroMark 2.2 reference space. If the relevance is lower, then the study results may be less likely to accurately translate, making them less comparable to other studies in the current review. The responses were recorded as follows: Yes = 1, No/Unclear = 0. The overall relevance of each study was determined based on the sum of all items: High Relevance = 5, Moderate Relevance = 3-4, Low Relevance = 0-2. A) Jensen, Calhoun, et al. (2024).

The objective of the current review was to identify a collection of data-driven whole-brain studies within recent rs-fMRI literature which examined group differences between individuals with schizophrenia or psychosis and controls and then translate their findings into a common reference space. The review focused on studies published within the last five years (2019-present) to capture studies published within approximately the same time frame since the NeuroMark approach was first developed and implemented, as comparability with the NeuroMark framework was an integral part of the review. By comparing findings across studies within the unifying framework of NeuroMark 2.2, the review aimed to inform the development of stable imaging markers of schizophrenia by determining which patterns of dysconnectivity were most consistent. The review also sought to compare potentially relevant features which may help to explain inconsistencies across studies.

## Methods

This systematic review was conducted according to the Preferred Reporting Items for Systematic reviews and Meta-Analyses (PRISMA; Page et al., 2021).

### Eligibility Criteria

The review sought to include primary empirical articles published in English in peer-reviewed journals in the last five years reporting analyses of human resting-state fMRI and utilizing cortical-subcortical-cerebellar terminology to describe schizophrenia or psychosis. All studies were required to include schizophrenia subjects in their clinical sample as that was the primary interest of the current review, however, other clinical profiles were included in the final article selection if they were part of a larger psychosis sample because much of the schizophrenia literature is confounded with psychosis. In addition, to maximize the comparability of findings, this review only included studies with data-driven case-control comparisons of whole-brain functional connectivity. The review sought to further refine its inclusion of data-driven approaches by only including studies which implemented ICA in delineating their regions of interest (ROIs).

### Information Sources, Search Strategy, and Data Extraction

Included articles were identified using PubMed, APA PsycInfo, and Google Scholar. The search was restricted to a five-year period spanning January 1, 2019 - February 4, 2025. Search terms included: (“schizophrenia” OR “psychosis”) AND “resting-state fMRI” AND (“cortical-subcortical-cerebellar” OR “cerebello-thalamo-cortical”). The titles and abstracts of 683 identified records were screened for basic criteria (see **Figure 1**). A secondary screening was performed, reviewing the method sections for the remaining 112 studies for analysis-specific requirements. Articles meeting all criteria were retained for final review. From each article, information was extracted pertaining to publication, acquisition details for the dataset(s) examined, demographics, clinical features, preprocessing, implementation of ICA, statistical approach, the results of analyses investigating associations between clinical variables and functional connectivity, and the results of case-control functional connectivity comparisons. To more accurately compare the results across studies within a unified framework, the results of the functional connectivity analyses were translated into the 14 subdomains from the NeuroMark 2.2 template (Jensen, Turner, et al., 2024) based on ROI descriptions provided within each article^1^. The assignment of each ROI to a corresponding NeuroMark subdomain was done manually by visually inspecting spatial maps, entering peak coordinates into Neurosynth (http://neurosynth.org/; Yarkoni et al., 2011), and considering the anatomical labels and descriptions provided in the article.

### Synthesis, Relevance, and Risk of Bias

The results of case-control functional connectivity comparisons were recorded for each pair of the 14 functional subdomains (e.g., Cerebellar (CB)-Occipitotemporal (OT)). Specifically, the directionality of case-control group differences was recorded with +1 representing hyperconnectivity, 0 representing no significant difference, −1 representing hypoconnectivity, and NA used to indicate that the given comparisons were not made^2^. Definitions of dysconnectivity differ across studies, however, to harmonize and aid in interpretation of the results in the current review, hyperconnectivity was defined as an increase in positive directionality (or a decrease in negative directionality) of FC in schizophrenia/psychosis relative to controls and hypoconnectivity was defined as an increase in negative directionality (or a decrease in positive directionality) of FC in schizophrenia/psychosis relative to controls. Static FC was used when available, but dynamic FC was used as an alternative if static FC was not available. When dynamic FC was used, results were recorded for all reported states and directionality was reported according to the same system if all states were in agreement, or based on the directionality of the sum of all states if they differed. A similar winner-take-all approach to recording was applied if the results of multiple datasets were reported separately or if the results within a given subdomain were mixed (e.g., some Cerebellar-Subcortical FC pairs are hyperconnected, while some are hypoconnected), in which case the directionality of significant results were tallied, and the result of the majority was recorded.

Considering the varying degree of similarity between ROIs across studies and the intrinsic connectivity networks (ICNs) defined in the NeuroMark atlas, and also the varying levels of detail articles include when describing ROIs, each study was evaluated to determine the relevance of its results when interpreted within this framework (see **Table 1**). Five questions were developed to assess the level of detail provided in describing ROIs (i.e, peak coordinates, spatial maps, anatomical labels), the comparability of the ROIs (e.g., a priori seed v. ICA-derived), and whether the functional connectivity analyses were comprehensive of the whole brain (e.g., comparisons between all pairs of ROIs). If the relevance was lower, then the study results may be less likely to accurately translate, making them less comparable to other studies in the current review. Responses to each were recorded as “Yes” (1) or “No/Unclear” (0). The overall relevance of the findings for each study were described by the sum of all items, with the possible total ranging from 0-5, with higher scores indicating that more criteria were met and therefore had higher relevance to the current review. For ease of interpretation, each study’s relevance was categorized as “High relevance” (score of 5), “Moderate Relevance” (scores ranging from 3-4), or “Low Relevance” (scores ranging from 0-2).

In addition, an optimized rating system was developed to assess risk of bias for each article in the current review (see **Table 2**). The JBI critical appraisal checklist for case control studies (https://jbi.global/critical-appraisal-tools), as well as risk of bias measures used in previous literature reviews (Ailion et al., 2017; Aleksonis & King, 2023; Steinberg & King, 2024) were referenced in developing an optimized rating system to assess each article identified for the current review. 13 questions were developed regarding study recruitment and inclusion criteria, the description, definition, and comparability of case/control samples, data acquisition and preprocessing, confounds addressed, and whether they corrected for multiple comparisons. Responses to each were recorded as “Yes” (1) or “No/Unclear” (0). For studies using datasets reported in previous studies (e.g., FBIRN) information provided previously (e.g., description of imaging acquisition) was considered. The overall bias assessment of each study was determined based on the sum of all items, with totals ranging from 0-13, with lower scores indicating that fewer criteria were met and therefore had a higher risk of bias. For ease of interpretation, each study’s risk of bias was categorized as “Low Risk” (scores ranging from 11-13), “Moderate Risk” (scores ranging from 7-10), or “High Risk” (scores ranging from 0-6).

**Table 2.**
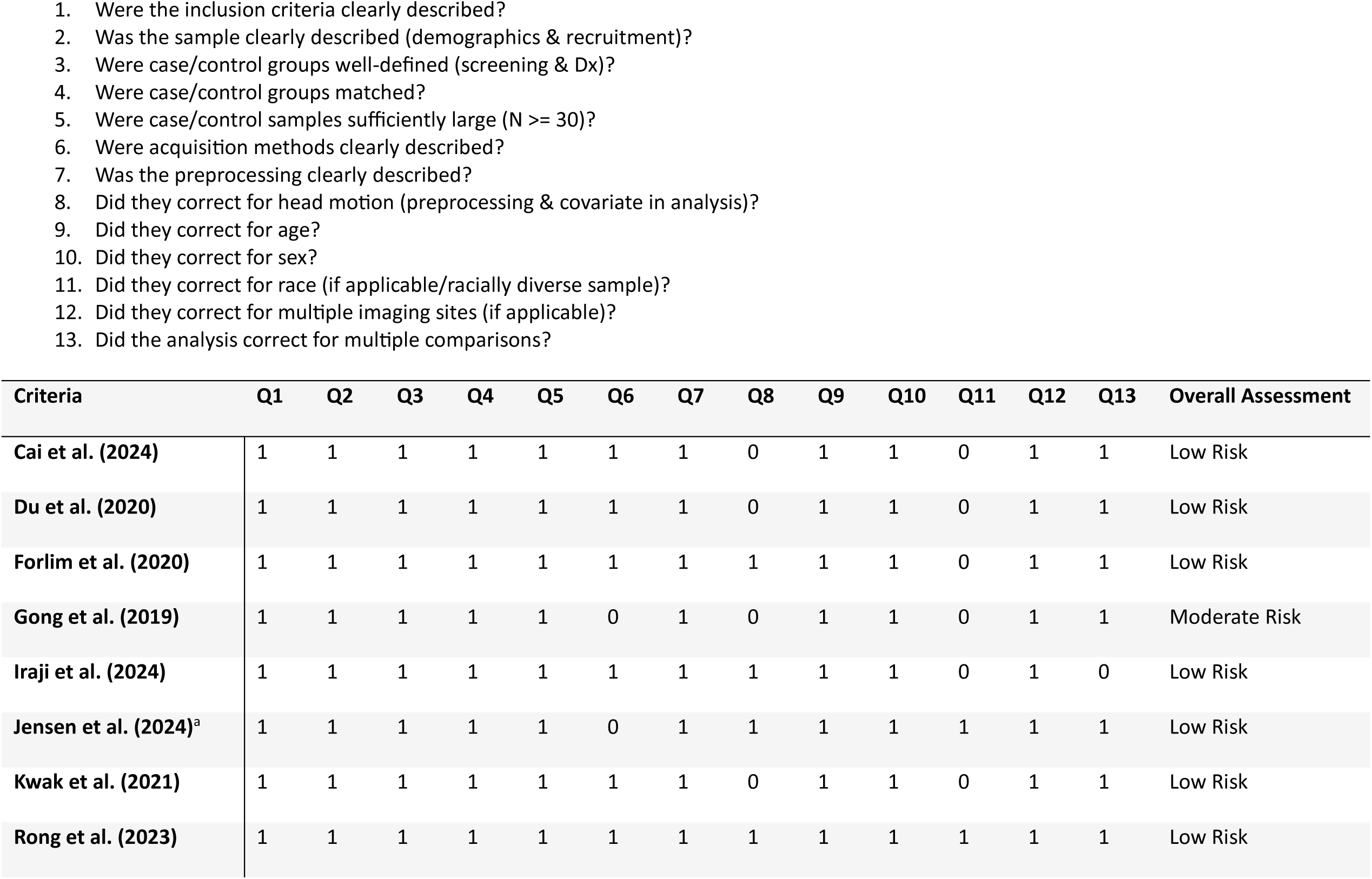
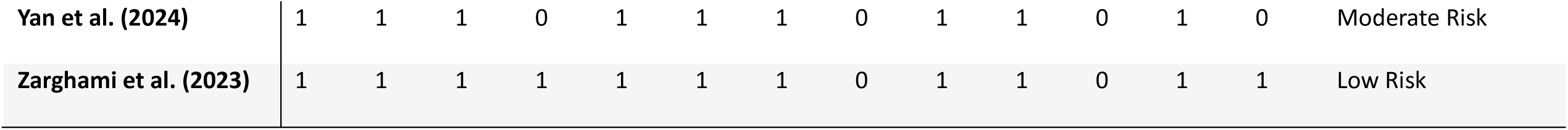
Risk of bias assessment. The risk of bias for each study was rated with the following criteria. The responses were recorded as follows: Yes = 1, No/Unclear = 0. For studies using datasets reported in previous studies (e.g., FBIRN), information provided in the original study was considered. The overall bias assessment of each study was determined based on the sum of all items: Low Risk = 11-13, Moderate Risk = 7-10, High Risk = 0-6. A) Jensen, Calhoun, et al. (2024).

## Results

### Study Selection and Sample Characteristics

Initial searches on PubMed, APA PsychInfo, and Google Scholar identified 683 records. After screening, 10 articles were determined to be eligible for inclusion in the final review. The reasons for exclusion are reported in **Figure 1**. Notably, 253 articles were excluded because they did not include a schizophrenia or psychosis sample (see **Figure 2**). Even within the final selection of articles, the sample types varied across studies (see **Table 3**), with two studies examining transdiagnostic early psychosis samples (Jensen, Calhoun, et al., 2024; Kwak et al., 2021), one study investigating comparisons with an early-onset schizophrenia sample (Cai et al., 2024), and a majority of seven studies investigating comparisons with chronic schizophrenia. Six studies utilized a single dataset, one study utilized two datasets but reported analyses of each separately (Du et al., 2020), one study utilized two datasets combined into one for its analyses (Jensen, Calhoun, et al., 2024), one study utilized three datasets combined (Iraji et al., 2024), and one study utilized five datasets combined (Yan et al., 2024). Four studies utilized overlapping datasets, although the inclusion of other datasets made each unique (Du et al., 2020; Iraji et al., 2024; Yan et al., 2024; Zarghami et al., 2023). The sample sizes ranged from 76 (35 schizophrenia) −2,615 (1302 schizophrenia). Cai et al. (2024) utilized an early-onset schizophrenia sample with a mean age of 15. The early psychosis studies examined slightly older samples with a mean age around 23 (Jensen, Calhoun, et al., 2024; Kwak et al., 2021). Rong et al. (2023) also utilized a relatively young chronic schizophrenia sample with a mean age of 25. The remaining six studies examined chronic schizophrenia samples with mean ages ranging from 35-39.

**Table 3.**
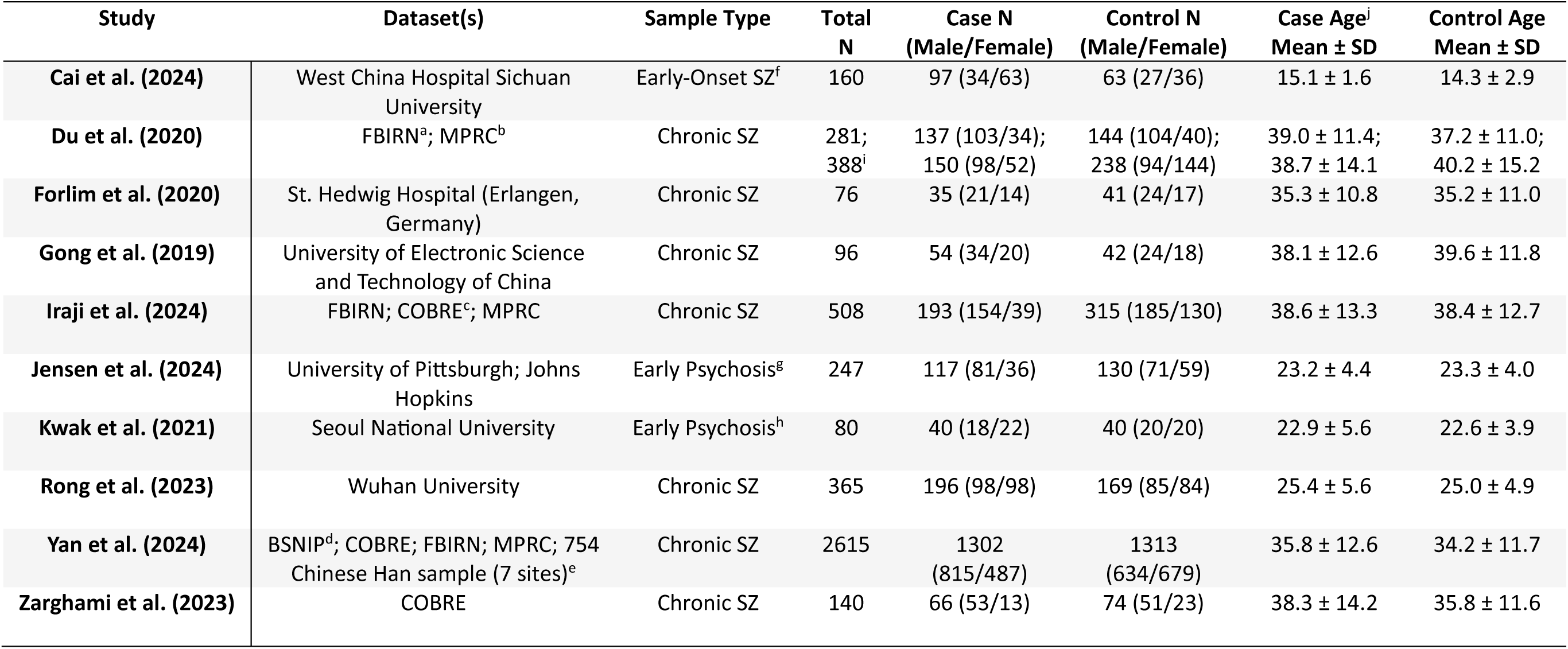
Demographic characteristics of the studies included in the review. A) FBIRN: Function Biomedical Informatics Research Network data repository (Keator et al., 2016). B) MPRC: Maryland Psychiatric Research Center (Adhikari et al., 2019). C) COBRE: Center for Biomedical Research Excellence (Aine et al., 2017). D) BSNIP: Bipolar-Schizophrenia Network on Intermediate Phenotypes (Tamminga et al., 2013). E) 754-Chinese Han Sample (7 sites): Peking University Sixth Hospital, Beijing Huilongguan Hospital, XinXiang Hospital Simens, Xinxiang Hospital GE, Xijing Hospital, Renmin Hospital of Wuhan University, Zhumadian Psychiatric Hospital (Yan et al., 2019). F) The early-onset schizophrenia sample consists of individuals diagnosed with schizophrenia before age 18. G) The early psychosis sample is transdiagnostic including individuals with schizophrenia, schizoaffective disorder, schizophreniform, bipolar disorder, major depressive disorder, and others within two years of first psychosis onset. H) The early psychosis sample is transdiagnostic including individuals with schizophrenia, schizophreniform, and schizoaffective disorder within two years of first psychosis onset. I) Demographics are reported separately for the two datasets in Du et al. (2020) because the analyses and results are reported for each dataset separately. J) Age is reported in years. SZ: schizophrenia

### Clinical Characteristics and Associations with FC

Clinical characteristics are summarized in **Table 4**. The clinical samples in all studies were diagnosed using DSM-IV or DSM-IV-TR criteria, except for the sample in Forlim et al. (2020) which was diagnosed based on ICD-10 criteria. Six of the studies utilized the positive and negative syndrome scale (PANSS; Kay et al., 1987) for schizophrenia, with four of them reporting associations between symptom severity and FC (Cai et al., 2024; Du et al., 2020; Gong et al., 2019; Rong et al., 2023). However, these findings were mixed, with no consistent patterns (see **Appendix 1**). The early-onset schizophrenia sample in Cai et al. (2024) had relatively high symptom severity with a mean PANSS total of 78.5 and was also characterized by a relatively short duration of illness (DOI < 1 year) and lower antipsychotic use (average chlorpromazine equivalence of 140.7 milligrams per day). Symptom severity was not as severe in the early psychosis samples (Jensen, Calhoun, et al., 2024; Kwak et al., 2021), and both had a much shorter DOI than the chronic schizophrenia samples. The sample reported in Jensen, Calhoun, et al. (2024) also had relatively low average antipsychotic use.

**Table 4.** Characteristics of the clinical samples included in the review. A) SANS and SAPS composite scores have been converted to PANSS Negative and Positive scores to make comparisons easier across studies (see van Erp et al., 2014). B) The mean PANSS total is not provided, however, it has been estimated as the sum of mean positive, negative, and general subscales (provided). C) SANS and SAPS global scores have been converted to PANSS Negative and Positive scores to make comparisons easier across studies. D) DOI: Duration of illness. E) CPZ: Chlorpromazine equivalence in milligrams per day. F-G) The olanzapine dose reported has been converted to CPZ based on Leucht et al. (2015). H) DM: Default Mode subdomain, ET: Extended Thalamic subdomain, FC: Functional Connectivity, CB: Cerebellar domain, SM: Sensorimotor domain, IT: Insular-Temporal subdomain, BG: Basal Ganglia subdomain, FR: Frontal subdomain, EH: Extended Hippocampal subdomain, OT: Occipitotemporal subdomain, OC: Occipital subdomain, TP: Temporoparietal subdomain, SA: Salience subdomain, PL: Paralimbic subdomain, SOC: Stockings of Cambridge from Cambridge Neuropsychological Test Automated Battery. SZ: schizophrenia

| Study | Diagnostic Criteria | Symptom Scale | Symptom Score Mean ± SD | DOI<sup>d</sup> Mean ± SD | CPZ<sup>e</sup> Mean ± SD | Findings Related to Symptom Severity, DOI, CPZ, & Cognitive Impairment<sup>h</sup> |
| --- | --- | --- | --- | --- | --- | --- |
| Cai et al. (2024) | DSM-IV | PANSS | 78.5 ± 24 PANSS total | 8.7 ± 13.2 months | 140.7 ± 210.5<sup>f</sup> | DM-ET FC positively correlated w/ PANSS general; SOC positively correlated with FR-ET FC (did not survive FDR) |
| Du et al. (2020) | DSM-IV-TR; DSM-IV | PANSS | --- | --- | --- | CB-ET FC negatively correlated w/ PANSS negative |
| Forlim et al. (2020) | ICD-10 | SANS; SAPS | ~13.9<sup>a</sup> PANSS Negative; ~15.1 PANSS Positive | 9.4 ± 8.8 years | 317.1 ± 221.6 | DM-DM FC negatively correlated w/ SANS composite and apathy; CPZ did not correlate w/ DM-DM FC, it also had no impact as a covariate |
| Gong et al. (2019) | DSM-IV | PANSS | ~63.2<sup>b</sup> PANSS total | 13.9 ± 10.6 years | 281.6 ± 122.7 | ET-SM FC positively correlated w/ PANSS total, PANSS general, & PANSS negative; ET-IT and ET-OT FC positively correlated w/ PANSS negative; ET-ET FC negatively correlated w/ PANSS negative; ET-ET FC negatively correlated w/ DOI; CPZ negatively correlated w/ ET-CB FC and positively correlated w/ OT-ET, SM-ET FC |
| Iraji et al. (2024) | DSM-IV-TR; DSM-IV-TR; DSM-IV | PANSS | 61.1 ± 15.4 PANSS total | --- | 344.8 ± 274.1 | CPZ did not correlate w/ IT/TP-SM, SA-DM FC |
| Jensen et al. (2024) | DSM-IV | SANS; SAPS | ~16<sup>c</sup> PANSS Negative; ~15 PANSS Positive | 11 (median) months | 210.5 ± 220.9 | Nothing survived FDR; SANS positively correlated w/ BG-FR FC, SM-EH FC; SANS negatively correlated w/ IT-OT FC, IT-FR FC, OC-FR FC, CB-CB FC; SAPS positively correlated w/ BG-FR FC; SAPS negatively correlated w/ ET-CB FC, EH-CB FC |
| Kwak et al. (2021) | DSM-IV | PANSS | 69.0 ± 14.3 PANSS total | 5.7 ± 3.8 months | 378.5 ± 296.0<sup>g</sup> | --- |
| Rong et al. (2023) | DSM-IV | PANSS; SANS; SAPS | 82.7 ± 11.6 PANSS total | 46.2 ± 54.1 months | 373.8 ± 283.7 | Nothing survived FDR; FR x SM FC positively correlated w/ PANSS total & negative; DM x DM FC negatively correlated w/ PANSS general; FR x SM FC positively correlated w/ PANSS positive; IT-SM FC negatively correlated with DOI; CPZ positively correlated w/ TP-SM FC |
| **Yan et al. (2024)** | DSM-IV; DSM-IV-TR; DSM-IV-TR; DSM-IV; DSM-IV-TR | --- | --- | --- | --- | --- |
| **Zarghami et al. (2023)** | DSM-IV-TR | --- | --- | --- | 363.1 ± 305 | All 7 MATRICS domains showed significant impairments in SZ; CCA top results were SM-SM and PL-TP EF, top MCCB traits were social cognition, reasoning/problem solving, working memory |

### Case-Control Group Differences in FC

The results of case-control functional connectivity for each pair of the 14 functional subdomains is summarized in **Figures 3** and **4**. **Figure 3a** displays the number of studies reporting hyperconnectivity across NeuroMark subdomains in schizophrenia/psychosis relative to controls, **Figure 3b** displays the number of studies reporting hypoconnectivity, and **Figure 3c** displays the net study count with hyperconnectivity represented as +1 and hypoconnectivity represented as −1. The most consistently reported patterns of dysconnectivity (net study count >= 5) are portrayed in **Figure 4**, with red arrows representing hyperconnectivity in schizophrenia/psychosis relative to controls and blue arrows representing hypoconnectivity. Although many notable patterns were observed, the most prevalent were cerebellar-cortical (CB-Sensorimotor (SM) & CB-Insular-Temporal (IT)) hyperconnectivity, cerebellar-subcortical (CB-Extended Thalamic (ET)) hypoconnectivity, subcortical-cortical (ET-SM, ET-OT, ET-Occipital (OC), ET-IT, Basal Ganglia (BG)-SM, BG-Temporoparietal (TP), & BG-IT) hyperconnectivity, and cortico-cortical (IT-OT) hypoconnectivity.

**Figure 3.**
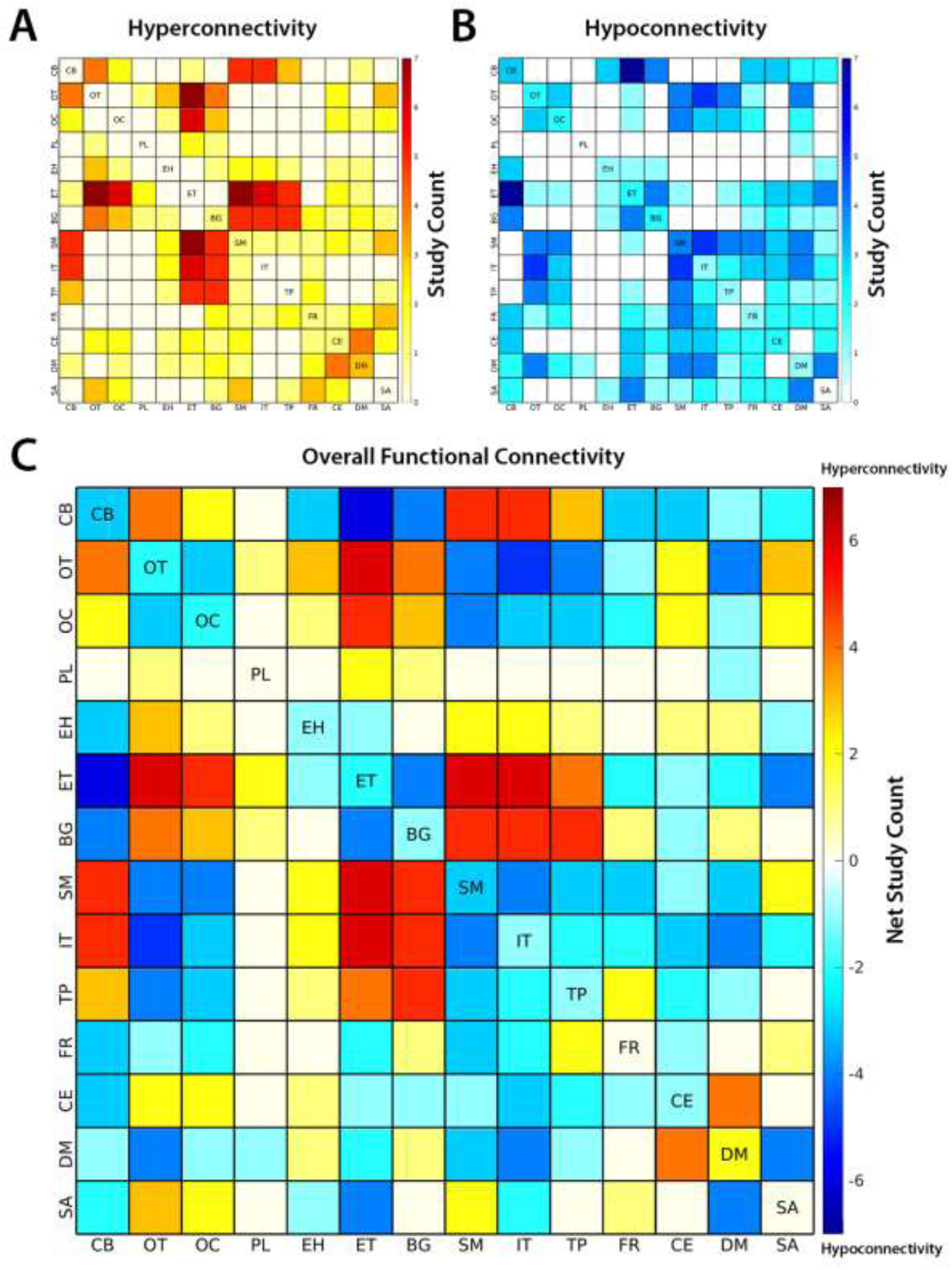
The number of studies reporting a) hyperconnectivity and b) hypoconnectivity between functional subdomains of the brain are shown above. Hyperconnectivity represents an increase in the positive directionality of functional connectivity (FC) in schizophrenia (SZ) relative to controls and hypoconnectivity represents a relative decrease (or increase in negative directionality) in SZ. The overall patterns of FC are represented by net study count, where studies reporting hyperconnectivity are assigned +1 and studies reporting hypoconnectivity are assigned −1. The 14 subdomains are based on the NeuroMark 2.2 multi-scale template: cerebellar (CB), visual-occipitotemporal (OT), visual-occipital (OC), paralimbic (PL), subcortical-extended hippocampal (EH), subcortical-extended thalamic (ET), subcortical-basal ganglia (BG), sensorimotor (SM), higher cognition-insular temporal (IT), higher cognition-temporoparietal (TP), higher cognition-frontal (FR), triple network-central executive (CE), triple network-default mode (DM), and triple network-salience (SA).

**Figure 4.**
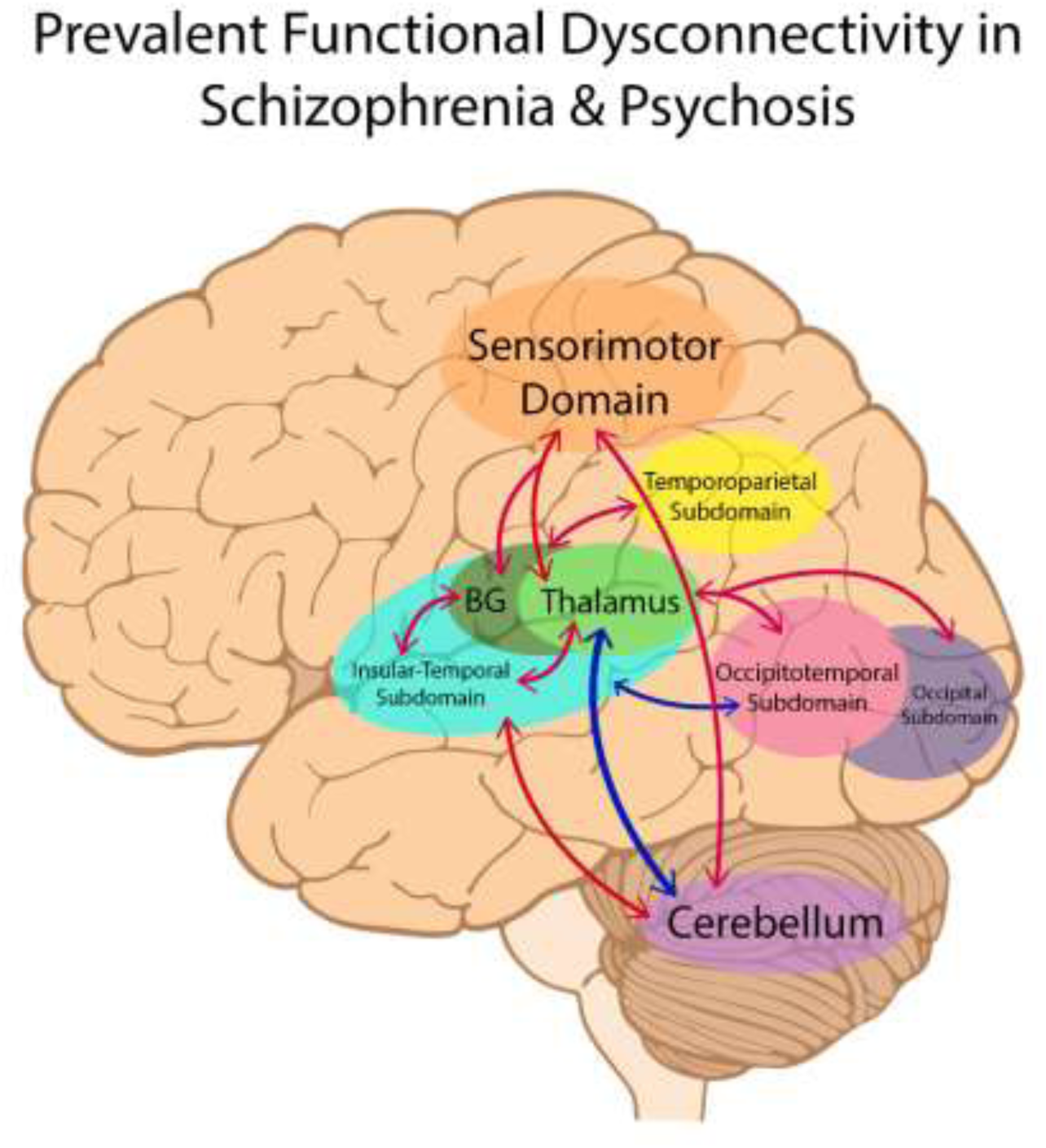
The most consistently reported patterns of dysconnectivity (with a net study count of 5 or more) are portrayed above. Hyperconnectivity in schizophrenia relative to controls is represented by red arrows between brain regions, with hypoconnectivity represented in blue. BG: Basal Ganglia

### Consistency, Risk of Bias, and Certainty of Evidence

To further evaluate the consistency of the FC case-control comparison results across studies, a Pearson correlation between the extracted FC results was calculated between studies (see **Figure 5**). While seven of the ten studies demonstrated relatively high similarity of results, Rong et al. (2023) displayed weaker correlations and Cai et al. (2024) and Forlim et al. (2020) appeared to be weakly anticorrelated with the common patterns.

**Figure 5.**
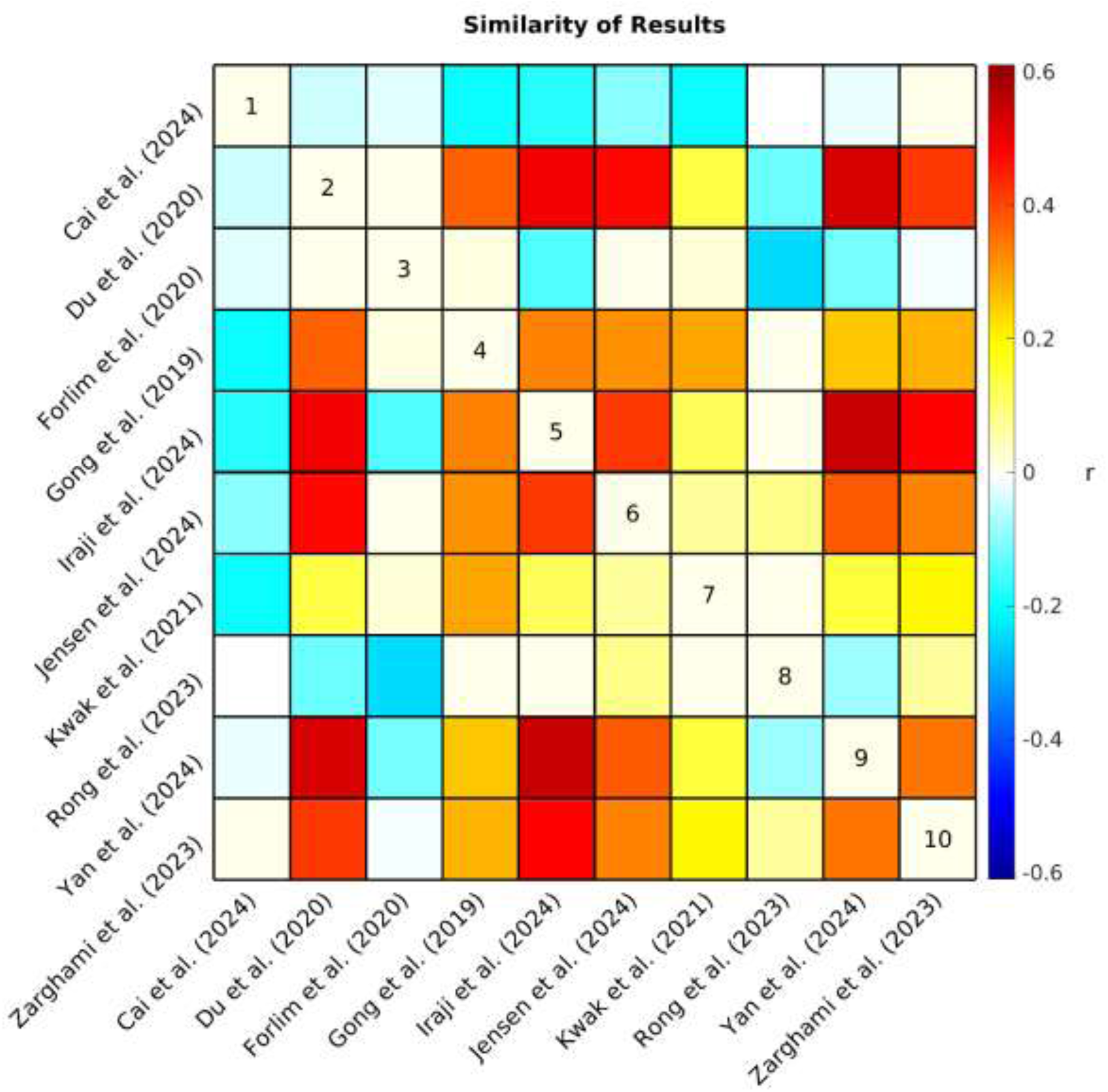
Each study produced an array of values (i.e., +1, 0, −1, NA) representing hyper/hypoconnectivity between case/control groups for each pair of subdomains. The similarity (Pearson correlation coefficient *r*) of observed patterns of functional connectivity across studies is represented by the correlation matrix above. NA values were interpreted as zero, therefore, a lower correlation suggests that studies had less similar results either because the observed patterns of connectivity differed between studies or because of differences in analytical approach (e.g., fewer comparisons were made due to differences in network definitions).

Eight of the ten studies were assessed as having low risk of bias, demonstrating that in general samples were carefully considered and appropriately matched, acquisition and preprocessing were clearly described, applicable confounds were controlled for, and analyses corrected for multiple comparisons. One notable exception is that only two studies (Jensen, Calhoun, et al., 2024; Rong et al., 2023) took race and ethnicity into consideration in their analyses. Two studies (Gong et al., 2019; Yan et al., 2024) were assessed as having moderate risk of bias, with Gong et al. (2019) penalized for lacking detail describing the procedures used for resting-state fMRI acquisition. In addition, although Gong et al. (2019) performed motion correction in preprocessing, they did not control for motion effects in their statistical analyses.Yan et al. (2024) likewise did not correct for motion effects in their basic group comparison of FC, nor did they correct for multiple comparisons as only schizophrenia minus control FC differences were provided (i.e., no test statistics were provided as these results were only supplementary to the primary analyses). In addition, it appears that the schizophrenia and control samples in Yan et al. (2024) were not matched by sex, although this was controlled for in the analysis.

The studies varied considerably in their implementation of ICA (see **Table 5**). The ROIs in three of the studies (Du et al., 2020; Jensen, Turner, et al., 2024; Yan et al., 2024) represented ICNs delineated through ICA spatially constrained to the NeuroMark 1.0 template (Du et al., 2020) which includes 53 optimized ICNs selected from a model order of 100. Although not identical, the ROIs utilized in Zarghami et al. (2023) were very similar, delineated through ICA spatially constrained to a 50 ICN template from Allen et al., (2014) which also used a model order of 100. Notably, these four studies were assessed as highly relevant (see **Table 1**) to the current review because they were evaluated to more accurately translate into the NeuroMark 2.2 reference space due to attributes of how the networks were delineated and described. Iraji et al. (2024) utilized the Group ICA of fMRI Toolbox (GIFT; http://trendscenter.org/software/gift) to perform group-level spatially-constrained ICA similar to the previous four studies, however, they utilized only 14 ICNs selected from a model order of 20. Lower model orders of ICA tend to be less granular which may limit the study’s ability to isolate effects and translate into the NeuroMark reference space which incorporates higher model orders (see Abou-Elseoud et al., 2010; Mirzaeian et al., 2024). Furthermore, this study was assessed as having moderate relevance, because the ICNs were described with limited detail, lowering the confidence in the accuracy of their assignment into NeuroMark 2.2 subdomains.

**Table 5.** Notes about the implementation of independent component analysis (ICA) in the functional connectivity analyses from each study included in the review. SZ: schizophrenia

| Study | Notes on Implementation of ICA | Model Order | Final ICNs | Labeling |
| --- | --- | --- | --- | --- |
| **Cai et al. (2024)** | ICA used to parcellate thalamus; FC represents thalamic subdivisions by whole-brain voxel-level analysis | --- | 5 thalamic ROIs, whole-brain voxel-level | "Yeo Network" and another not specified for labeling "Regions" |
| **Du et al. (2020)** | ICA spatially-constrained to NeuroMark 1.0 | 100 | 53 | NeuroMark 1.0 |
| **Forlim et al. (2020)** | Used GIFT/ICA to extract ICNs, visually inspected and labeled ICNs, then compared within network connectivity between case/control; did not examine between network FC and did not include CB ICNs | 21 | main analysis included DMN only, but exploratory included additional 6 ICNs | "templates in GIFT" & "experts" |
| **Gong et al. (2019)** | ICA used to parcellate thalamus; FC represents thalamic subdivisions by whole-brain voxel-level analysis | --- | 6 thalamic ROIs, whole-brain voxel-level | --- |
| **Iraji et al. (2024)** | Used GIFT/ICA to extract ICNs, visually inspected and labeled 14 ICNs, 4 states of dynamic FNC reported between all 14 ICNs | 20 | 14 | Previous knowledge/studies |
| **Jensen et al. (2024)** | ICA spatially-constrained to NeuroMark 1.0 | 100 | 53 | NeuroMark 1.0 |
| **Kwak et al. (2021)** | ICA used on thalamus by whole-brain seed-based correlation maps to identify 20 thalamus-related ICNs, FC between thalamus and network ROIs | 20 | 20 | Based on networks reported by Yuan et al. (2016) |
| **Rong et al. (2023)** | HCP ICA templates, 32 ROIs (avg of all voxels within seed) from 8 ICA derived RSNs, FC between 32x32 ROIs, no subcortical/thalamic ROIs | --- | ICA templates used as mask for ROIs | HCP ICA templates |
| **Yan et al. (2024)** | ICA spatially-constrained to NeuroMark 1.0, reported difference between SZ-control (but no t-stats/p-vals) | 100 | 53 | NeuroMark 1.0 |
| **Zarghami et al. (2023)** | ICA spatially-constrained to Allen et al. (2014) 50 ICN template | 100 | 50 | Allen et al. (2014) |

Forlim et al. (2020) differed more in their approach, using GIFT to perform blind ICA with a model order of 21. They also differed in that they selected only a single ICN to represent the DMN, which was the focus of their primary analyses (i.e., DMN-DMN FC). They also performed limited exploratory analyses with an additional six ICNs they selected from the 21. In both primary and exploratory analyses, however, the ICNs were used to perform cluster-based analyses which differed from the whole-brain FC comparisons made in the previous five studies which compared the correlation between the time courses of other ICNs. This study was assessed as having moderate relevance to the current review due to these key differences in analytical approach. Rong et al. (2023) also differed in that it used ICA-derived templates to select 32 ROIs. The FC from these ROIs was calculated from the average of all voxels within each seed rather than correlations between ICN time courses. Furthermore, they did not include any subcortical ROIs in their analyses and were assessed as having only moderate relevance to the current review. The remaining three studies (Cai et al., 2024; Gong et al., 2019; Kwak et al., 2021) differed considerably from the previously mentioned studies in that ICA was performed only on the thalamus. Specifically, Kwak et al. (2021) identified 20 thalamus-related network ROIs spanning across the brain and examined seed-based functional connectivity between them. The relevance of this study was limited (assessed as moderate) due to a limited description of these networks as well as limited comparisons made between these networks. Cai et al. (2024) and Gong et al. (2019) used ICA to parcellate the thalamus into five and six thalamic ROIs respectively. In both studies, FC represented the correlation between thalamic subdivisions and whole-brain voxel-level analyses. Due to this limitation, Gong et al. (2019) was assessed as having only moderate relevance. Cai et al. (2024) was further limited to low relevance as spatial maps were not provided for all the relevant ROIs, making it more difficult to accurately translate the results into the NeuroMark reference space.

## Discussion

### Cortical-Subcortical-Cerebellar Terminology

Many articles in the initial search were excluded because they did not include a schizophrenia or psychosis sample (see **Figure 2**). Movement disorders were the largest contributor, composing 122 of these articles. Interestingly, the initial search identified nearly as many studies examining samples with movement disorders as those examining schizophrenia or psychosis. It is possible that this finding may be due to overlapping affected brain circuitry between schizophrenia and movement disorders (Andreasen et al., 1998). Indeed, some of the most prevalent patterns reported in the current review were dysconnectivity in motor pathways (see **Figure 4**). Regardless of the reason, it is apparent that not only is the term “cortical-subcortical-cerebellar” frequently used to describe a prominent category of pathology unrelated to schizophrenia and psychosis, but it is also used to describe a wide range of disorders and even non-clinical samples. This undermines the usefulness of the term for characterizing the biological profiles of schizophrenia and psychosis and further demonstrates the need for more precise language in defining the key alterations in rs-fMRI in individuals with these clinical profiles.

### Aberrant Connectivity in Schizophrenia and Psychosis

The primary objective of the current review was to delineate and describe in greater detail the most prevalent patterns of dysconnectivity observed in schizophrenia and psychosis with data-driven ICA approaches. The most prominent patterns observed in schizophrenia and psychosis can be summarized as hypoconnectivity between cerebellar and subcortical structures, specifically between the cerebellum and thalamus, with hyperconnectivity between cerebellar and cortical structures (SM & IT) as well as between subcortical (ET & BG) and cortical structures (SM, TP, IT, OT, & OC). Cortical-cortical connectivity (e.g., IT-OT) was less commonly reported, but when it was the patterns typically reflected hypoconnectivity in schizophrenia and psychosis.

Consistent with Andreasen and colleague’s (1998) theory of cognitive dysmetria and the cerebello-thalamo-cortical framework (Harikumar et al., 2023; W. J. Hwang et al., 2022), the cerebellum and thalamus appear to be key nodes, and disruptions in circuitry between them and the cortex appear to be characteristic of schizophrenia and psychosis. Indeed, cerebellocortical and thalamocortical dysconnectivity have been suggested as a substrate of under-regulated cognitive processes (Andreasen et al., 1999; Clark et al., 2020). Notably, the most consistent patterns of cortical dysconnectivity were centered in regions associated with primary and secondary sensory and motor processing. Specifically, the SM spans across primary somatosensory and motor cortex as well as related heteromodal association areas (Jensen, Turner, et al., 2024). Motor-related symptoms such as tardive dyskinesia (TD) are often experienced in schizophrenia and disruptions in sensorimotor circuitry are widely implicated across studies (Andreasen et al., 1998; Cattarinussi et al., 2023; Walther et al., 2017). Neurological abnormalities in sensorimotor performance are sometimes referred to as neurological soft signs and have been suggested to be present during the early stages of schizophrenia and psychosis and potentially even pre-date illness onset (Dazzan & Murray, 2002). However, it remains unclear to what extent TD and alterations in SM can be attributed to the pathophysiology of schizophrenia or to the effects of antipsychotic medication (Lerner & Miodownik, 2011; Yu et al., 2021). Indeed, Cai et al. (2024) reported ET-SM hypoconnectivity in contrast to the ET-SM hyperconnectivity reported by the majority. Importantly, the sample in Cai et al. (2024) had the lowest antipsychotic dosage as well as a relatively short DOI. That being said, Jensen, Calhoun, et al. (2024) reported ET-SM hyperconnectivity in a sample with only a small increase in dosage and duration. Unfortunately, only four of the reviewed studies specifically tested for associations between medication and FC, with two implicating SM circuitry (Gong et al., 2019; Rong et al., 2023) and two reporting null results (Forlim et al., 2020; Iraji et al., 2024; see **Appendix 1**). Consistent with these findings, Gong et al. (2019) and Rong et al. (2023) also identified associations between SM circuitry and DOI (**Appendix 1**), which is generally associated with increased medication use. Further investigation is warranted to disentangle the specific effects of medication and chronicity on sensorimotor FC.

Sensory association areas within the SM, along with the IT and TP, encompass key cortical regions implicated in auditory and language networks. Specifically, primary auditory cortex (A1) is located within the medial superior temporal lobe (a region with high spatial overlap with the IT) and the dorsal auditory stream leads from A1 to the parietal lobe (overlapping with the TP and SM; Jensen, Turner, et al., 2024; Purves et al., 2001; Rauschecker & Tian, 2000). Prior studies have attributed auditory verbal hallucinations (AVHs) to disruptions in the processing of auditory information across these networks (Jardri et al., 2011; Kompus et al., 2011; Kuhn & Gallinat, 2012). Similarly, the OT and OC encompass cortical regions primarily associated with visual sensory processing, including the primary visual cortex (V1), visual association cortex (V2-V5), and the ventral visual stream (Goodale & Milner, 1992; Huff et al., 2024; Jensen, Turner, et al., 2024). Like AVHs and auditory cortex, visual hallucinations (VHs) have been attributed to disruptions in processing in visual cortex, although the underlying mechanisms remain unclear (Collerton et al., 2023).

### Addressing Heterogenous FC Across Studies

There was a consensus in many of the findings across studies and although some of this may be driven partly by similarities in datasets and analytical approach, these factors cannot fully account for the consistent findings, as there were also many differences. For example, among the seven studies showing higher similarity of results, three of them included subjects from the FBIRN and MPRC datasets (Du et al., 2020; Iraji et al., 2024; Yan et al., 2024) and three included subjects from the COBRE dataset (Iraji et al., 2024; Yan et al., 2024; Zarghami et al., 2023), however, different combinations of these datasets enabled all of these studies to examine FC group differences in unique samples. Furthermore, Gong et al. (2019), Jensen, Calhoun, et al. (2024), and Kwak et al. (2021) demonstrated high similarity with these studies despite their use of completely different datasets. On a similar note, Jensen, Calhoun, et al. (2024) and Kwak et al. (2021) both utilized early psychosis samples with mixed diagnostic groups as opposed to the schizophrenia samples utilized by the other eight studies, however, it appears that neither reduced chronicity, nor the inclusion of additional diagnostic groups made a substantial difference, as these two studies reflected relatively high similarity with most of the other studies. Although, further investigation into the effects of chronicity may be warranted when considering the divergent results of the early-onset schizophrenia sample observed in Cai et al. (2024), as well as the associations between DOI and dysconnectivity reported by Gong et al. (2019) and Rong et al. (2023).

While differences in analytical approach likely contributed to differences in results across studies, this cannot completely account for the observed differences in FC and in some cases these methodological differences may have had minimal impact. For example, four of the studies were highly similar in their implementation of ICA (Du et al., 2020; Jensen, Calhoun, et al., 2024; Yan et al., 2024; Zarghami et al., 2023), with three of them defining ROIs as ICNs which were spatially constrained to the same standardized network template (Du et al., 2020; Jensen, Calhoun, et al., 2024; Yan et al., 2024). However, some of the other studies showing high similarity of results had fairly large differences in analytical approach, for example, Iraji et al. (2024) examined dynamic FC in 14 ICNs selected from a model order of 20 but was highly similar to many of the studies utilizing a model order of 100 to examine static FC, with especially high similarity to Yan et al. (2024). Similarly, Kwak et al. (2021) utilized a model order of 20 to examine thalamus by whole-brain FC and showed relatively high similarity with the studies utilizing ICNs from higher model orders. Gong et al. (2019) differed even more, using ICA only in its parcellation of thalamic subdivisions which were used as seeds in a whole-brain voxel-level analysis, yet demonstrated fairly high similarity of results when compared with the six studies using only ICA-derived ICNs as ROIs. Together, these findings suggest that the differences in results were not merely a product of the differences in samples and analytical approach but instead were likely driven by additional factors as well.

One possibility is that the divergent studies may be capturing different biological profiles of schizophrenia. Schizophrenia is likely biologically heterogenous and different samples may present different combinations of biologically different subgroups, displaying mixed results (Andrés-Camazón et al., 2025; Clementz et al., 2016; Feczko et al., 2019). There is evidence for this in the varying clinical profiles of the samples examined in Rong et al. (2023), Cai et al. (2024), and Forlim et al. (2020; see **Table 4**), the three studies displaying low similarity with the majority (see **Figure 5**). Specifically, Rong et al. (2023) observed the highest symptom severity among the reviewed samples. Similarly, Cai et al. (2024) examined FC in an early-onset schizophrenia sample displaying higher symptom severity than most of the reviewed samples. Early-onset schizophrenia has previously been reported to have a different clinical profile than adult-onset schizophrenia (Immonen et al., 2017; Remschmidt & Theisen, 2012), which appears to be largely what the other studies are capturing in their samples although their participants are not explicitly described that way. Indeed, early-onset (also referred to as childhood or adolescent onset) schizophrenia has been suggested to reflect a different biological profile as well, with differences reported in genetic associations (Alkelai et al., 2023) and functional connectivity (Zhang et al., 2021). A study examining age-related changes in FC even observed opposite patterns in FC directionality between younger and older psychosis cohorts (Passiatore et al., 2023). Similarly, Anticevic et al. (2015) observed hyperconnectivity in prefrontal cortex regions where hypoconnectivity is more commonly observed and suggested that some patterns of aberrant FC may be inverted in early-course schizophrenia, reflecting dynamic alterations in FC as individuals transition to chronic schizophrenia, possibly resulting from compensatory mechanisms. It is possible that age-related factors such as those described in prior work may help account for the anticorrelated results observed between Cai et al. (2024) and majority of the findings included in the current review. Future studies are needed to substantiate this potential reversal of dysconnectivity between early and adult-onset cohorts.

In contrast to Cai et al. (2024) and Rong et al. (2023), the sample in Forlim et al. (2020) appeared to have relatively low symptom severity, although differences in reported scales makes it difficult to compare across studies (see van Erp et al., 2014). In addition, Forlim et al. (2020) utilized ICD-10 which differs from the diagnostic criteria of the DSM-IV and DSM-IV-TR which was utilized by the other nine studies. Although generally considered to be comparable, it has been suggested that the ICD-10 results in a broader concept of schizophrenia than the DSM-IV (Cheniaux et al., 2009; Lindström et al., 1997). Forlim et al. (2020) also had the smallest sample size with only 35 individuals with schizophrenia. Together, these factors may have resulted in a more unique clinical sample. Although the samples in Rong et al. (2023), Cai et al. (2024), and Forlim et al. (2020) all share the diagnostic label of schizophrenia, each appears to represent a unique clinical profile which may have contributed to its divergent patterns of FC. The field would benefit from approaches which leverage biological measures to further explore the possibility of unique signatures of FC among subtypes within psychosis spectrum disorders such as schizophrenia (Ballem et al., 2025).

The differences in terminology used to describe functional units across studies should also be considered. As previously mentioned, variability in terminology or inconsistencies in how the same terminology is applied to different functional entities (Uddin et al., 2023), as well as individual variability in their spatial maps (Jensen, Turner, et al., 2024) may potentially confound our ability to interpret and compare results between studies. Furthermore, differences in each study’s implementation of ICA, such as higher or lower model order impacts key characteristics of the derived ICNs, such as their granularity (Mirzaeian et al., 2024). As model order increases, functional units tend to branch out into smaller units (Abou-Elseoud et al., 2010). It is possible that FC between some ICNs derived from lower model orders which are spatially large, spanning multiple networks, may tend to zero out the effects of smaller regions within them as they average across a larger area and move towards a global baseline. For example, in Rong et al. (2023), OT and OC networks were combined into a single visual network, potentially weakening the observed effects and contributing to the null results observed in the visual network, which was an area with some of the most consistently reported aberrations across studies. Similarly, in Kwak et al. (2021), the cerebellum, temporal lobe, and parietal lobe were all represented by a single component, potentially confounding some of the cerebellum-related results with those of cortical regions. These inconsistencies in functional entities make it difficult to interpret results within a common framework and highlight one of the limitations of the current review. Indeed, Cai et al. (2024) displayed the greatest inconsistency with other studies in the analysis and was also assessed as having the lowest relevance to the current study due to more ambiguous reporting of ROIs as well as significant variations in its methodological approach. Similarly, Forlim et al. (2020) and Rong et al. (2023) were rated as having only moderate relevance to the current study because they made relatively limited comparisons which neglected cerebellar (Forlim et al., 2020) and subcortical (Rong et al., 2023) regions in their analyses. It is plausible that the low similarity of findings in these three studies is largely reflective of a lack of information due to their less comprehensive analyses. This observation may underscore the importance of future studies focusing efforts on more detailed reporting of their ROIs as well as employing more comprehensive data-driven whole-brain approaches.

### Insights, Limitations, and Recommendations

The current review also yielded many valuable insights into different subdomains within the brain. For example, the paralimbic subdomain (PL) was the subdomain with the most null results across studies, primarily because relatively few studies (Cai et al., 2024; Gong et al., 2019; Kwak et al., 2021; Zarghami et al., 2023) reported ROIs which overlapped with these regions. Notably, the spatial area covered by the PL is relatively small and may require the increased granularity offered by higher model orders, possibly explaining why it is less commonly delineated in existing literature. In contrast to the PL, there were some networks, such as those within the triple network domain (TN) and frontal subdomain (FR), which were frequently implicated across studies and yet also seemed to yield a lower net study count (see **Figure 4**) due to inconsistent findings. One possibility is that networks incorporating anterior association cortex, such as these, are more frequently involved in higher cognitive processes and are more likely to vary across individuals and studies than more primal networks like motor and sensory cortex (e.g., visual, auditory, & somatosensory; Mueller et al., 2013; Sun et al., 2022). Indeed, higher cognitive networks incorporating the prefrontal cortex, such as those within TN and FR, may be more dynamic, with FC varying more over time (Iraji et al., 2019). One limitation of studies examining static FC, is that they may not capture the full range of variability of more dynamic brain networks, and as a result, findings may be less consistent for these networks across studies. Iraji et al. (2024) in the current review may lend support to this notion, as the observed group differences for the CE (or frontoparietal/attention networks) changed across different states. Unfortunately, one limitation of the current review is that it was not sensitive to time-varying changes such as this, but instead was constrained to comparisons of static FC. As dynamic FC approaches are more widely employed to capture these patterns, the field would greatly benefit from a standardized approach to summarizing and comparing dynamic FC across studies.

## Conclusions

Andreasen et al. (1998) and many others (Anticevic et al., 2015; W. J. Hwang et al., 2022; Menon et al., 2023; Woodward & Heckers, 2016; Zhou et al., 2015) have focused on the prefrontal cortex as a key node in the cognitive dysmetria framework, viewing schizophrenia as a disease of higher cognitive functions. While there is much evidence supporting dysconnectivity in this node and the implicated etiology is feasible, findings are largely inconsistent across studies. This is likely because the complex relationship between this node and others requires more sophisticated analytical approaches, for example dynamic FC approaches which are sensitive to time-varying changes. Therefore, although the prefrontal cortex remains integral to understanding the neurobiological substrate of schizophrenia and psychosis, concentrating on this node may not be effective for establishing stable imaging markers, at least while employing analytical approaches which investigate static FC in rs-fMRI. Instead, adapting new analytical strategies, or focusing on nodes in the cerebellum, thalamus, and primary motor and sensory (e.g., SM, IT, & OC) or possibly more posterior association cortex (e.g., TP & OT) may prove to be a more effective approach. Further investigation is needed to explore how these patterns of dysconnectivity vary in relation to medication and chronicity as well as across individuals with unique clinical profiles within schizophrenia and psychosis spectrum disorders.

## Author Contributions

KMJ contributed through conceptualization, methodology, software, formal analysis, investigation, data curation, writing – original draft, writing – review & editing, and visualization. TZK contributed through conceptualization, writing – review & editing, and supervision. PAC contributed through writing – review & editing. VDC contributed through conceptualization, writing – review & editing, resources, supervision, and funding acquisition. AI contributed through conceptualization, writing – review and editing, and supervision.

## Data Availability

All data produced in the present study are available upon reasonable request to the authors.

https://trendscenter.org/data/

## Appendix 1 Clinical Associations with FC

Symptom associations with FC were mixed, with Cai et al. (2024) reporting a positive correlation between DM-ET FC and PANSS general, Du et al. (2020) reporting a negative correlation between CB-ET FC and PANSS negative, Gong et al. (2019) reporting a positive correlation between SM-ET FC and PANSS total, PANSS general, and PANSS negative, and a positive correlation between IT-ET and OT-ET FC and PANSS negative, and a negative correlation between ET-ET FC and PANSS negative. Although results did not survive FDR correction, Rong et al. (2023) reported a positive correlation between FR-SM FC and PANSS total, PANSS positive, and PANSS negative and a negative correlation between DM-DM FC and PANSS general. Three studies utilized the scale for the assessment of negative symptoms (SANS; Andreasen, 1983) and the scale for the assessment of positive symptoms (SAPS; Andreasen, 1984), although only two reported associations between symptom severity and FC (Forlim et al., 2020; Jensen, Calhoun, et al., 2024). Forlim et al. (2020) reported a negative correlation between DM-DM FC and SANS composite score as well as the individual item for apathy. Although none of the symptom associations survived FDR correction, Jensen, Calhoun, et al. (2024) reported positive correlations between SANS global and FR-BG FC and SM-EH FC, as well as between SAPS global and FR-BG FC. Jensen, Calhoun, et al. (2024) also reported negative correlations between SANS global and IT-OT FC, IT-FR FC, OC-FR FC, and CB-CB FC, as well as between SAPS global and CB-ET FC and CB-EH FC.

Only two studies tested for associations between duration of illness (DOI) and FC, with Gong et al. (2019) reporting a negative correlation between DOI and inter-thalamic FC and Rong et al. (2023) reporting a negative correlation between DOI and IT-SM FC. Four studies tested for associations between antipsychotic use and FC, with Gong et al. (2019) reporting a negative correlation between CPZ and CB-ET FC and a positive correlation between CPZ and OT-ET FC and SM-ET FC, and Rong et al. (2023) reporting a positive correlation between CPZ and TP-SM FC. Forlim et al. (2020) and Iraji et al. (2024) tested for correlations between CPZ and FC but reported no significant relationships.

Only two studies examined associations between cognitive assessments and FC. Cai et al. (2024) reported a positive correlation between Stockings of Cambridge and FR-ET FC, although this association did not survive FDR correction. Zarghami et al., (2023) reported significant impairments in schizophrenia across all seven MATRICS domains with top CCA results of SM-SM EF and PL-TP EF with top traits of social cognition, reasoning/problem-solving, and working memory.

## Footnotes

1 Referenced articles were utilized when pre-existing templates were employed.

2 Fewer FC pairs are possible in studies constrained to larger spatial scales, for example, extended thalamic subdomain (ET) – basal ganglia subdomain (BG) FC cannot be calculated if both of those subdomains are represented by a single ROI.

